# Protocol for a pragmatic non-randomized controlled study evaluating the efficacy of a dimensional adaptation of GPM (GPM-extended) compared to a classic outpatient GPM treatment for patient fulfilling criteria for BPD: the EPSYLIA project

**DOI:** 10.1101/2025.04.09.25325532

**Authors:** Martin Blay, Inès Benmakhlouf, Marion Zenou, Mélissa Amate, Arjin Uludag-Martin, François Chevrier, Marie-Laure Pomey, Marie Brisebarre, Paul Lebourleux, Oyhana Lopepe, Saioa Lagadec, Loïs Choi-Kain, Mario Speranza

## Abstract

**Background:** Personality disorders (PD) are common and debilitating psychiatric conditions, often characterized by severe interpersonal and self-dysfunction. Borderline personality disorder (BPD) is the most studied PD, with treatments like Good Psychiatric Management (GPM) demonstrating effectiveness. To address the current state of the personality disorder field, where most evidence-based treatments have been developed using a categorical approach, while the most empirically supported approach is the dimensional one, we developed an adaptation of GPM (GPM-extended) integrating concepts of dimensional personality dysfunction into an established therapeutic framework.

**Methods:** This prospective “here-there” study will compare GPM-extended with standard GPM in two outpatient facilities. Participants (≥18 years) meeting BPD criteria per the SCID-II will be included. The GPM-extended group incorporates a dimensional diagnostic framework focusing on three personality dilemmas (rejection/abandonment fears, self-esteem dysregulation, and perfection/control issues), with tailored psychoeducation and treatment priorities. The standard GPM group will follow the validated GPM protocol, emphasizing interpersonal hypersensitivity and standard psychoeducation. Both groups will receive weekly individual therapy and group interventions for one year. The primary objective is to assess the efficacy of GPM-extended in reducing BPD symptoms after one year. Secondary outcomes include personality functioning, traits, and various clinical dimensions such as impulsivity, emotional regulation, and social functioning.

**Discussion:** This study seeks to evaluate the feasibility and efficacy of integrating dimensionality into GPM, offering a pragmatic pathway to improve PD treatment and bridging gaps between evidence-based conceptualizations and treatments of PD.

**Clinical trial number:** NCT06913738

## 1. Introduction

Personality disorders (PD) are among the most common psychiatric disorders. They affect around 10% of the general population, and the proportion of patients with at least one personality disorder reaches 50% in psychiatric settings (1). These disorders are associated with an increased risk for somatic comorbidities (e.g. (2)), psychiatric comorbidities (e.g., (3)), with a fewer remission rate (e.g., (4)) and a fewer efficacy of their treatment (e.g., (5)). Moreover, these disorders are associated with large consequences in term of social and professional functioning (6,7) and in terms of societal costs (8,9). Finally, personality disorders are also associated with stigmatization, both in general population and among health care professionals (10).

The most studied personality disorder is borderline personality disorder (BPD). BPD affects around 1.6% of the general population and is characterized by hypersensitivity to rejection and abandonment, resulting in significant difficulties in interpersonal relationships, emotion regulation and identity (11). Since its inclusion in the DSM-III (12), the BPD diagnosis has allowed the development of a large amount of research that shed light on the importance to consider, diagnose, and possibly treat personality disorders (11,13), that were often considered as rigid, chronic and less important disorders compared to main disorders like major depressive or bipolar disorders (14). Indeed, major longitudinal studies have shown that BPD could improve over time (6), and evidence-based effective treatments have been developed over the last decades, each linked with a specific treatment conceptualization and framework, and having the overall same efficacy (15). The main therapies actually recommended are dialectical behavioral therapy (DBT), transference-focused psychotherapy, mentalization-based therapy (MBT) and schema-focused therapy (16). To note, these therapies are the most developed currently in the entire personality disorders field, as very few models have been developed for other PD categories. However, as these specialized models developed, an issue quickly emerged from a public health point of view, namely, the lack of accessibility and availability of these treatments for most BPD patients. Thus, given that these therapies are highly specialized and poorly available (17), to overcome this issue clinicians have developed new models, less complex and specialized but more accessible and practicable. These models are based on what is thought to be the core characteristics of effective treatments for borderline patients, and have been popularized through the dissemination of two mains models: Good Psychiatric Management (GPM) and Structured Clinical Management (18).

However, despite this robust clinical and scientific foundation supporting the diagnosis of BPD, ongoing debates persist about its reliability, along with that of other traditional personality disorder categories. Indeed, these diagnoses are often the target of critics (19,20), because they are thought to have low reliability, with high level of diagnostic comorbidity (most patients have often more than one PD) and within-disorder heterogeneity (patients with the same PD can have different clinical issues) (14,19). In this context, researchers have developed the so-called “dimensional” models, describing personality disorders using the combination of two criteria: level of personality functioning (criteria A) and personality traits (criteria B). The two main models currently used are the Alternative Model of Personality Disorders of the DSM-5 (AMPD) (21) and the personality disorders model of the International Classification of Disease 11th version (ICD-11) (22). More precisely, criteria A is divided in two main dimensions, self and interpersonal functioning, and criteria B is composed of 5 personality traits, mostly derived from the Big-5 literature: negative affectivity, antagonism, disinhibition, detachment, and psychoticism (for the AMPD) / anankastia (for the ICD-11)). However, these new models are currently only scarcely used in clinical services (23). Moreover, they are also not free of critics, particularly with regard to the clinical relevance of personality traits, considered by some authors to not represent the complex and dynamic self- and interpersonal processes characteristics of PD patients (24). Our hypothesis is that these two issues may be linked, as clinicians may be more reticent to use new models when the latter are not sufficient to describe what they see in their daily practice. Thus, these models need to be enhanced, possibly using more comprehensive and relevant conceptualizations of PDs patients’ daily issues, which may help the transition from classifications to clinical practice. Such adaptation could be even more important given that these models have not been used in the development and assessment of evidence-based treatments (contrary to classic categorical models), which furthermore limits their usefulness, even if new models are being developed trying to fill this gap (25).

We believe that GPM could be useful to solve this issue, both in terms of diagnostic assessment and treatment. We discussed the diagnostic assessment aspect in a recently published case report (24). Briefly, GPM, whether we consider the BPD model or its adaptation for narcissistic (NPD) (26) and obsessive-compulsive personality disorders (OCPD) (27), has allowed the development of dynamic and fundamentally interpersonal models aiming to better describe the borderline (*interpersonal hypersensitivity* model), narcissistic (*intrapsychic coherence* model), or obsessive-compulsive (*model of overcontrol*) patient’ psychopathology than classic categories or personality traits. Notably, these three conceptualizations encompass the main dilemmas an individual with PD can face in his/her pathology: fear of rejection and abandonment (for BPD), self-esteem dysregulation (for NPD), and control dependency (for OCPD). Based on these assumptions, we previously described how incorporating these three conceptualizations by describing them as non-exclusive ways personality can dysfunction in one individual, rather than restricting them to one category, may be useful to offer a more relatable conceptualization than classic personality traits, while being a more dimensional and less rigid way of assessing personality disorders (24).

However, the contribution of GPM may not be limited only to diagnostic assessment part but may also encompass the therapeutic dimension. Recent research have shown that BPD criteria may be a relevant marker of personality functioning, even if questions remain about whether this applies to specific BPD criteria, all criteria collectively, or the overall severity of BPD symptomatology (28,29) Thus, if personality dysfunction is a core aspect of every personality disorder, and if BPD criteria are relevant markers of personality dysfunction, one could legitimately infer that all personality disorders may be treated by partly the same psychotherapeutic content, i.e., one developed and shown useful to treat patient fulfilling criteria for BPD, like GPM (30,31). This makes even more sense when considering the improvement in interpersonal functioning – one hallmark of personality functioning - in GPM trials, and the overall focus on getting a life outside of treatment to build an integrated sense of identity – another hallmark of personality functioning - in the GPM treatment framework (32). Finally, even if the three adaptations of GPM consider different personality disorders and are partly different, a large part of the content remains the same: use of diagnostic disclosure, psychoeducation, case management, progress, psychodynamic and behavioral treatment, multimodality, safety planning, symptomatic medication management… This makes a dimensional GPM adaptation feasible beside being theoretically useful, with 1°) this common part being the cornerstone of the adaptation, and 2°) the specific parts of each dilemma being provided depending on the patient’s profile.

Altogether, like classic dimensional models, GPM offers the possibility for dimensional approach, with personality functioning assessed with the presence/absence of BPD criteria, and characteristics of self- and interpersonal dysfunction considered globally using GPM’s dilemma-based approach. However, unlike classic dimensional models, GPM has been empirically tested and found efficient to treat patients fulfilling criteria for BPD, and provides a more simple, easy-to-understand, psychoeducation-accessible, and daily-experience related approach of personality characteristics than classic dimensional models. In this context, we designed a dimensional adaptation of GPM (*GPM-extended*) for patients fulfilling criteria for BPD. In the present paper, we will describe the protocol of a pragmatic, real-life based, “here-there” study assessing the effectiveness of an outpatient treatment program based on GPM-extended compared to an outpatient treatment program based on classic GPM.

## 2. Protocol

### 2.1. Design and settings

Our study will use a “here-there” methodology, i.e., the inclusion of patients will be made based on which facility each patient is referred to. Two facilities will be included, one offering the outpatient GPM-extended program (ADDIPSY, Lyon, France), and the other offering the outpatient classic GPM program as it has been validated in the literature (Clinique CARADOC, Bayonne, France). Both facilities are private day-hospitals, offering outpatient programs for a wide range of psychiatric disorders. 2 psychiatrists and 3 psychologists (in ADDIPSY), and 3 psychiatrists and 3 psychologists (in CARADOC) will take part as therapists in the current study. All the psychometric evaluation, data collection and entry will be conducted by two trained clinical research assistants (IB and SL).

The estimated inclusion period for the present study is five years. Recruitment started on June 19^th^ 2024 and is expected to be completed by June 2029. Each participant’s involvement lasts for one year, determined by the intervention period, which was established based on research standards for studying the effectiveness of psychotherapies in BPD. Data collection is anticipated to be completed by December 2029. The final results are expected to be available for publication by the end of 2030. Total duration of the study is estimated at six years and six months. However, the study may conclude earlier if the required sample size is reached ahead of schedule. The full SPIRIT schedule of enrolment, interventions, and assessments can be found in **Figure 1**.

**Figure 1.**
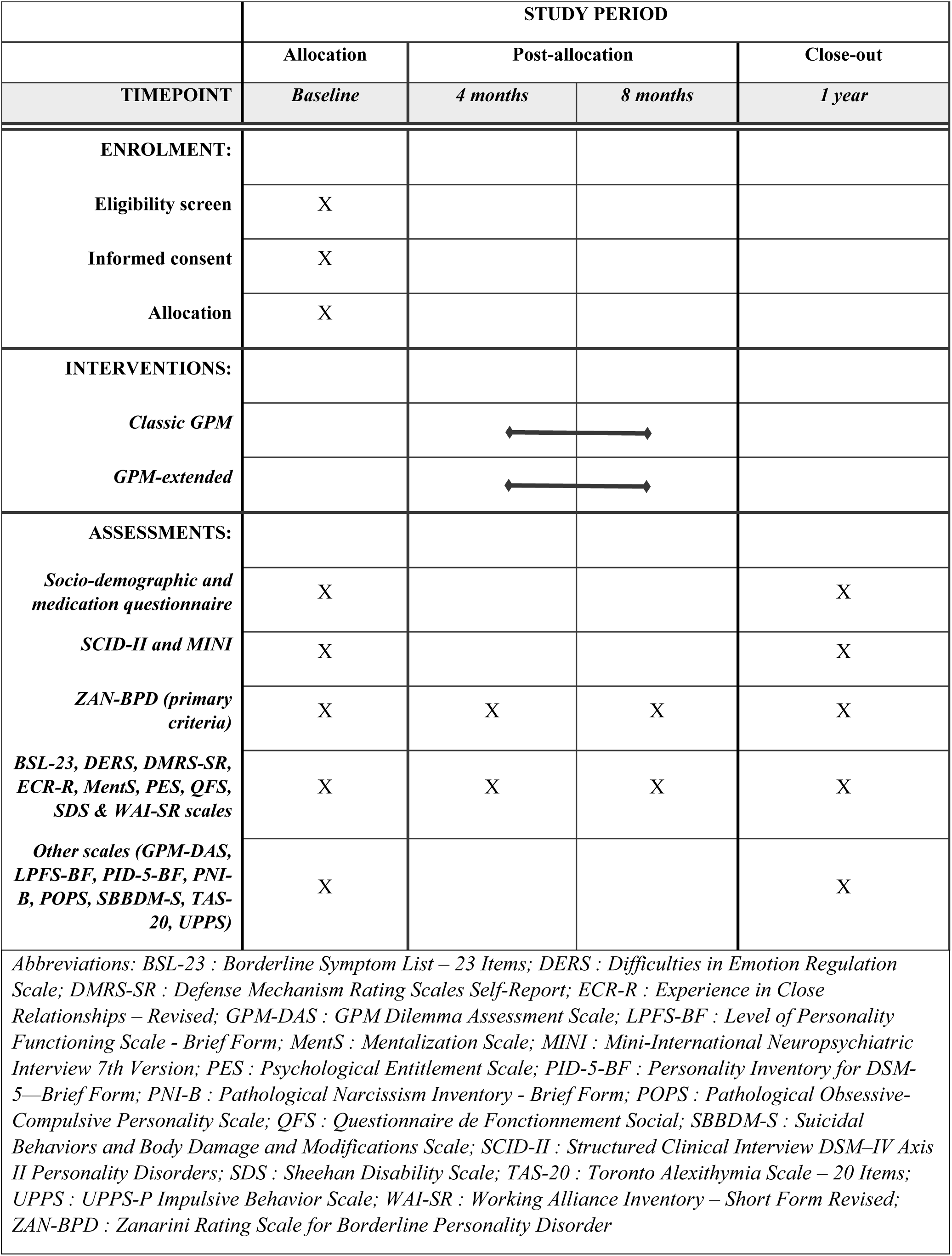
SPIRIT schedule of enrolment, interventions, and assessments.

Written informed consent will be gathered for each patient by the clinical research assistants and the referent psychiatrists in the facilities. The study protocol was submitted and validated by the Comité de Protection des Personnes Ile de France III (N°ID-RCB = 2024-A00475-42). To, even though this study meets the WHO definition of a clinical trial, it was not registered before the inclusions began because the authors were unaware that registration was required. The study has now been registered retrospectively on ClinicalTrials.gov under the identifier NCT06913738. The authors confirm that all ongoing and related trials for this intervention are registered. Finally, all substantial modifications to the protocol (e.g., changes in eligibility criteria, outcomes, or analytical strategy) will be submitted by the coordinating investigator to the sponsor’s representative for approval. Following a favorable opinion, the sponsor will initiate the necessary regulatory procedures to obtain approval from the relevant ethics committee and/or competent authorities. Relevant parties, including investigators, regulatory bodies, trial registries, and journals, will be informed of approved amendments as required.

### 2.2. Objectives and hypotheses

The primary objective of this study will be to evaluate the efficacy of GPM-extended compared to classic GPM in improving the overall level of BPD symptoms in patients meeting the criteria for the latter. Our secondary objectives will be to compare the efficacy on other dimensions of interests, including sub-dimensions of BPD (i.e., affective, cognitive, impulsive, interpersonal), personality functioning and intensity of personality traits (according to the alternative DSM-5 model), level of disability and social functioning, narcissistic and obsessive-compulsive symptomatology.

Overall, we expect an improvement in all the outcomes in both groups, notably in borderline symptoms and level of personality functioning, at 1 year of treatment. We also expect a significant difference in both groups, with GPM-extended being superior to classic GPM, notably on borderline, narcissistic, obsessive-compulsive symptoms and overall personality functioning.

### 2.3. Selection criteria

The inclusion criteria for the present study will be 1°) being >18 years old, 2°) having a diagnosis of BPD made using the Structured Clinical Interview DSM–IV Axis II Personality Disorders (SCID-II, (33)), 3°) having signed an informed consent and 4°) being affiliated with or beneficiary of the French social security system. On the other hand, exclusion criteria will be 1°) being < 18 years, 2°) having a comorbid psychotic disorder, intellectual deficiency, severe antisocial features, major substance use disorder making intensive psychotherapy impossible, anorexia nervosa with somatic risk, bipolar disorder in acute manic phase, 3°) being under protective measure, 4°) being unable to cooperate or complete self- or hetero-questionnaire and 5°) subject not affiliated with or non-beneficiary of a French social security system.

### 2.4. Key features of GPM-extended

Given that the present paper aims to present the protocol of our study, we will not describe all the characteristics and detail of our GPM-extended adaptation. A GPM-extended manual has been written for clinicians participating in the present study and is available upon request to the corresponding author. In this paragraph, we will rather discuss the specificity of our adaptation compared to classic GPM treatment (32).

GPM-extended builds on the foundational principles of GPM for BPD by incorporating a more dimensional and pragmatic approach. It emphasizes diagnostic clarity, treatment prioritization, personalized psychoeducation, and multimodal interventions tailored to severe personality dysfunctions. The diagnostic process focuses on BPD criteria and integrates dimensional tools like the Level of Personality Functioning Scale – Brief Form (LPFS-BF) (34) to assess personality functioning. This process is enhanced using the three main dilemmas a patient with severe personality dysfunction may face according to GPM (i.e., fear of rejection and abandonment, self-esteem dysregulation, and control dependency), to assess *how* one’s personality may dysfunction. The assessment of the presence/absence and relevance of each dilemma for each patient relies on both clinical and psychometric investigation, associated with repeated team discussion in the weekly supervision sessions. Clinical investigation uses DSM’s Section II and Section III specific criteria, and psychometric investigation uses the newly developed GPM Dilemmas Assessment Scale (GPM-DAS, **Table 1**) as well as other specific psychometric scales (i.e., Zanarini Rating Scale for Borderline Personality Disorder (ZAN-BPD), Pathological Narcissism Inventory Brief Form (PNI-B) and Pathological Obsessive-Compulsive Personality Scale (POPS). The complete dilemma assessment process can be found in **Table 2**.

**Table 1.**
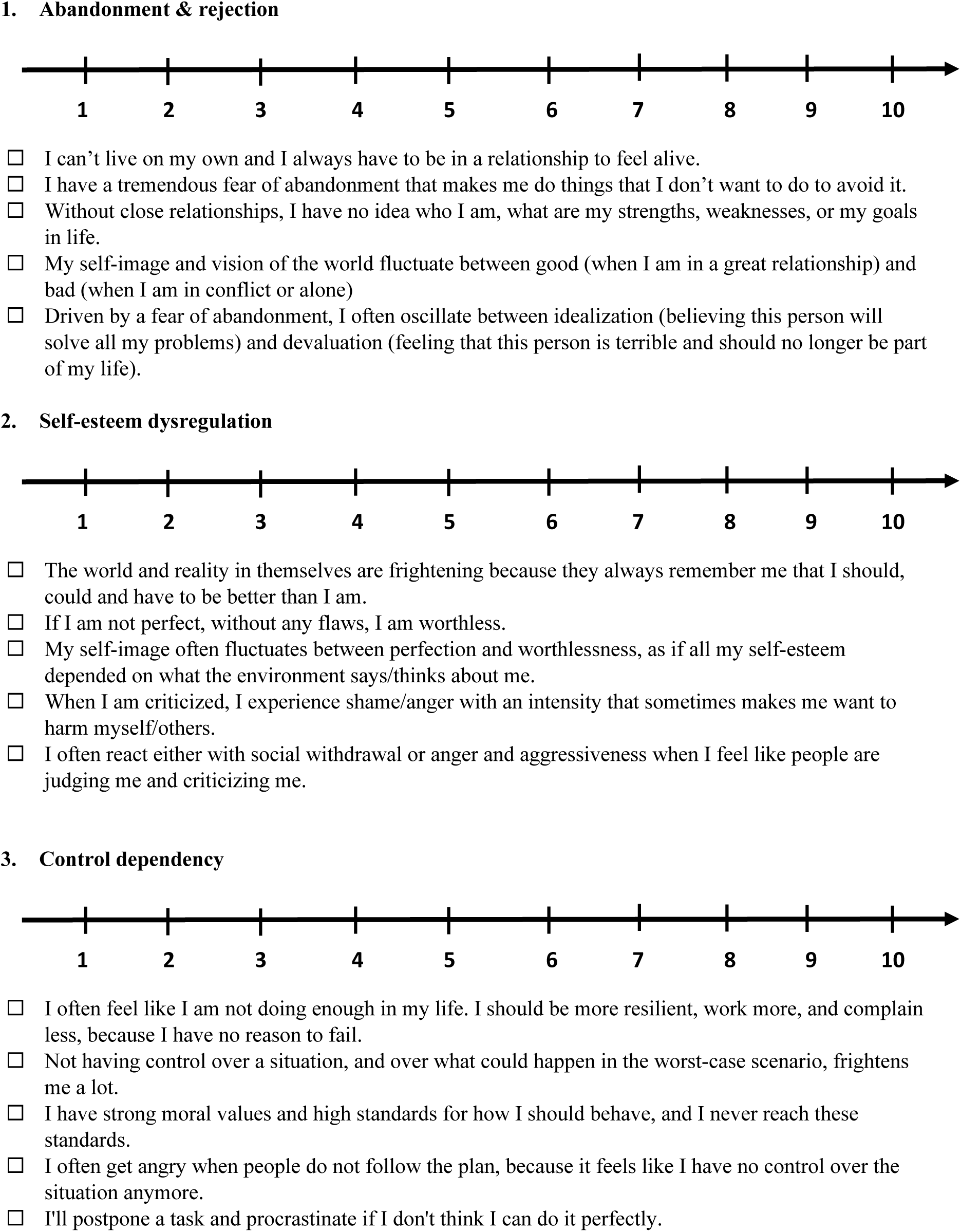
Content of the GPM-Dilemmas Assessment Scale (GPM-DAS) This questionnaire aims to identify what are the main triggers/dilemmas you are facing as a patient suffering from personality disorder. Here, you’ll find three scales rating from 0 (“not at all”) to 10 (“completely true”) that will help us adapt the treatment to what suits best for you. You’ll also find several examples of everyday situation to help you rate the scale. Please rate the intensity **over the last month**. There are no right or wrong answers, so please do your best.

**Table 2.** Dilemma assessment process.

| Table 2. Dilemma assessment process |  |  |  |
| --- | --- | --- | --- |
| Dilemmas (GPM models) | Clinical* | GPM-DAS | Other psychometric tools |
| Fear of abandonment & rejection (*interpersonal hypersensitivity*) | - Frantic efforts to avoid real or imagined abandonment<br>- Pattern of unstable and intense interpersonal relationships characterized by alternating between extremes of idealization and devaluation.<br>- Relationships marked by mistrust (&) neediness | For every dilemma:<br>- Non-relevant: 0-2<br>- Mildly relevant: 3-5<br>- Moderately relevant: 6-8<br>- Strongly relevant: 9-10 | ZAN-BPD 6th (abandonment) and 9<sup>th</sup> criteria (idealization and devalorization) >= 2 (moderate) |
| Self-esteem dysregulation (*intrapsychic coherence model*) | - Excessive reference to others [...] for self-esteem regulation; exaggerated self-appraisal [...] vacillating between extremes<br>- Emotion regulation mirrors fluctuations in self-esteem<br>- Relationships [...] exist to serve self-esteem regulation |  | PNI-B grandiosity and/vulnerability scores >=3<sup>#</sup> |
| Control dependency (*model of overcontrol*) | - Sense of self derived predominantly from work or productivity<br>- Rigid and unreasonably high and inflexible standards of behaviors; overly conscientious and moralistic attitudes<br>- Perfectionism that interferes with task completion |  | POPS total score >= 180<sup>#</sup> |
| * Only the three most relevant criteria are listed. |  |  |  |
| # The thresholds have been chosen subjectively based on the mean scores found in general population in the validation studies. |  |  |  |

Once the main dilemmas are identified, both the clinician and the patient work on a clear prioritization of the treatment targets, with the most impacting dilemmas being the first one to focus on. Indeed, GPM-extended tailors psychoeducation and case management to each patient’s specificities. This prioritization is often reassessed by the patient and the clinician throughout the treatment, to ensure that treatment addresses the most impactful issues while maintaining flexibility to adapt to evolving needs.

Regarding treatment content, GPM-extended retains the core principles of GPM, such as active support, validation, focus on real-life situations, and fostering patient responsibility for change (32). It also retains the core components of a GPM treatment (i.e., alliance building, goal setting and achievement, anticipation of difficulties, and narrative work), and emphasizes the importance of group interventions (supportive group, psychoeducation, or specialized approaches like DBT or MBT) and pragmatic pharmacological management. However, it expands upon these characteristics with a dimensional approach that emphasizes individualized treatment focusing on the patient’s dominant personality dilemmas identified in the diagnostic process. Indeed, alliance building challenges, narrative work, and treatment main targets are different depending on the dilemmas. For example, in the case of borderline symptomatology, treatment focuses on building a meaningful life (e.g., stable social and professional roles) over intense relationships; in the case of narcissistic symptomatology, treatment focuses on the development of an integrated sense of self-worth by addressing grandiosity, shame, and avoidance challenges; and, in the case of obsessive-compulsive symptomatology, treatment focuses on improving control dependency, perfectionism, and rigid standards and fears through emotional and behavioral corrective experiences.

### 2.5. Interventions

Both treatment programs have a 1-year length, as in most standard clinical studies on psychotherapy in BPD. Patients that require it will be offered a second year of treatment if it is clinically relevant. Patients refusing to participate in the present study will be treated just as the participants but will not undergo the protocol-related psychometric evaluation. A schematic representation of the study and programs content can be found in **Figure 2**. To note, given that both participating facilities provide a wide range of general psychiatric care (including individual and group therapies, such as trauma-focused or addiction-focused treatments), participants are allowed to engage in these additional interventions, as long as they are not specifically designed to target borderline personality disorder or related personality dysfunction.

**Figure 2.**
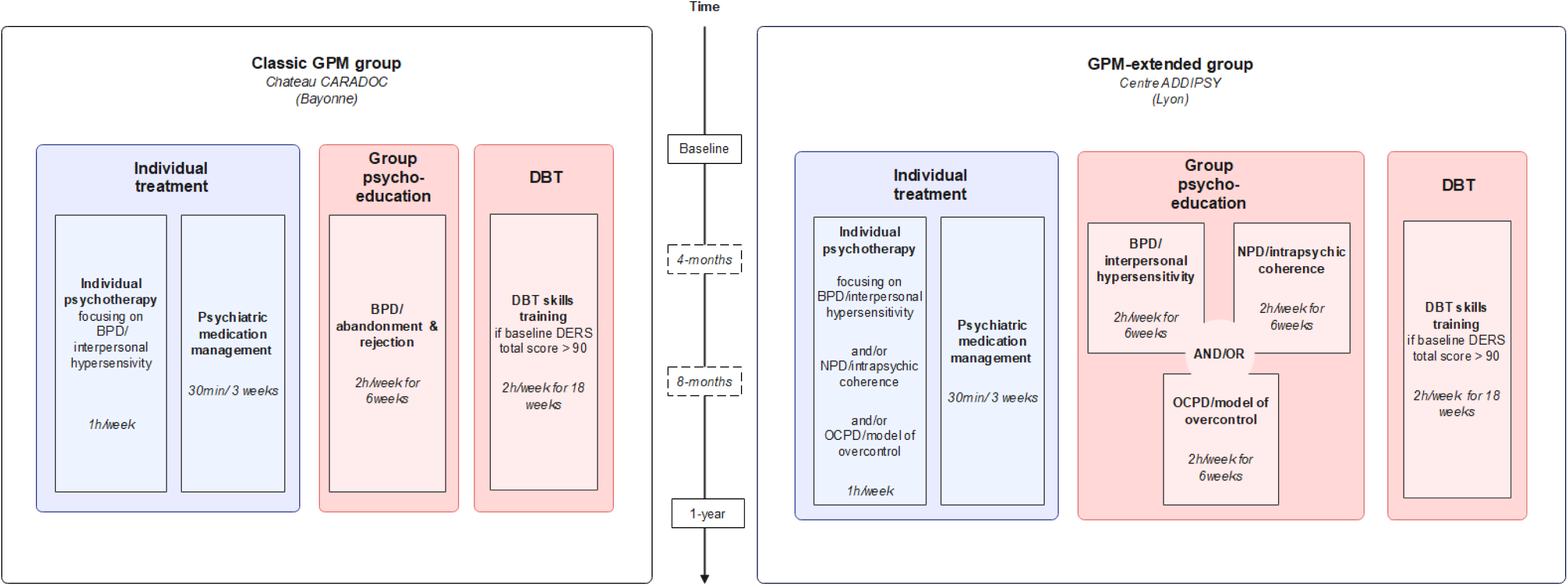
Schematic representation of the study groups.

#### Classic GPM group

The outpatient classic GPM group will consist in the association of both individual *and* group therapy:

- Individual = 1°) 1h weekly individual psychotherapeutic session conducted by a therapist trained in GPM, with a focus on interpersonal hypersensitivity *and* 2°) 30 min every 3 weeks of an individual psychiatric case management focused on medication stabilization by a psychiatrist trained in GPM.
- Group = 1°) Alongside the beginning of therapy, every patient will undergo >= 1 session of a 6-weeks group psychoeducation on BPD and 2°) Patients with high levels of emotion dysregulation (diagnosed when baseline Difficulties in Emotion Regulation Scale (DERS) total score is higher than the mean scores obtained in clinical populations, i.e. > 90 (Hallion et al., 2018)) will undergo >= 1 session of a 18-weeks DBT skills training group (either in one block or sub-divised in 3 blocks of 6 weeks each, corresponding to the main modules of DBT – mindfulness/distress tolerance, interpersonal efficacy and emotion regulation) during the year of therapy. To note, the exact moment when patients will be included will depend on the availability of skills groups).

The team will be trained and supervised (1 hour every week – online – and 1 day every month – in person) throughout the process by the main investigator of the study (MB). The latter will not practice as a therapist in the control group and will only serve as trainer and supervisor. All the content in this control group is manualized and validated, including the BPD psychoeducation program (35) and the DBT skills training program (36). Moreover, every clinician will be provided with the Handbook of Good Psychiatric Management for BPD for the conduction of individual psychotherapy (32).

#### GPM-extended group

The outpatient GPM-extended group will also consist in the association of both individual *and* group therapy:

- Individual = 1°) 1h weekly individual psychotherapeutic sessions conducted by a therapist trained in GPM, with a specialized focus on the main triggers/dilemmas identified with the patient *and* 2°) 30 min every 3 weeks of an individual psychiatric case management focused on medication stabilization by a psychiatrist trained in GPM.
- Group = 1°) Alongside the beginning of therapy, every patient will undergo >= 1 session of one or several of the 6-weeks group psychoeducation programs, depending on the main dilemmas of the patient (borderline, narcissistic and/or obsessive-compulsive, with each patient undergoing from 1 to 3 different programs, depending on the results at the GPM-DAS) *and* 2°) Patients with high levels of emotion dysregulation (diagnosed when baseline DERS-F total score is higher than the mean scores obtained in clinical populations, i.e. > 90 (Hallion et al., 2018)) will undergo >= 1 session of a 18-weeks DBT skills training group (either in one block or sub-divised in 3 blocks of 6 weeks each, corresponding to the main modules of DBT) during the year of therapy. Once again, the exact moment when patients will be included will depend on the availability of skills groups.

The team will be trained and supervised (1 hour every week – in person) throughout the process by the main investigator of the study (MB). The latter will practice as a therapist in the active group and will also be the trainer and supervisor. Regarding the content, a GPM-extended manual will be provided for clinicians, including the specific content of therapy for each core triggers/dilemmas. These core triggers/dilemmas will be identified with the patient using both classic psychometric tools (e.g., PNI-B, POPS) and our newly developed GPM-DAS, designed to assess the three main dilemmas considered in GPM models. Moreover, the BPD psychoeducation program and the DBT skills training program will be the same as those used in the “classic GPM” group. The psychoeducation program for NPD is also manualized and has recently been published (37), and the psychoeducation program for OCPD has also been published online recently (38).

### 2.6. Measures

#### Psychometric assessment

Every measure will be assessed by dedicated clinical research assistants. Included patients will be administered, at the inclusion and 1 year follow-up several semi-structured interview and self-reported questionnaires, including socio-demographic characteristics and prescribed treatment information. Moreover, included patients will also undertake several self-report questionnaires at 4 and 8 months of therapy, for further mediation analyses (cf. 3.6). All the different measurements carried out at each stage of the EPSYLIA project can be found in Figure 1.

##### Clinician-rated assessment

To assess categorical personality disorders, we will use the SCID-II (33,39). The SCID-II has great inter-rater reliability and internal consistency (40,41). Criteria will be rated if pathological, pervasive, and persistent, and we will use the recommended thresholds to establish diagnoses (e.g., for BPD, >= 5 out 9 criteria).

To assess other psychiatric disorders, we will use the Mini-international neuropsychiatric interview 7^th^ version (MINI) (42). The MINI is a valid and reliable tools, with good psychometric properties (43), that assesses a wide range of past and present neuropsychiatric disorders, including depressive, bipolar, psychotic, anxiety, obsessive-compulsive, post-traumatic stress, substance use and eating disorders.

Finally, to have a clinician-rated measure of BPD symptoms intensity as a primary outcome, we will use the ZAN-BPD (44). Overall, the ZAN-BPD has good internal consistency (Cronbach’s α=0.85). It is a 9-items scale used to assess each of the nine criteria for BPD on a five-point anchored rating scale of 0 to 4. A total score (sum of all items) and 4 sub-scores (affective, cognitive, impulsive, and interpersonal (35)) can be calculated.

##### Self-report personality assessment tools

To assess GPM dilemmas, we will use the GPM-DAS), which was specifically developed for this study. The GPM-DAS is an interviewer-rated 3-items scale, with each item assessing the presence or relevance of one dilemma— fear of rejection and abandonment, self-esteem dysregulation, or control dependency —on a 10-point Likert scale, over the last month. Each item is accompanied by a list of several examples illustrating how the dilemma may manifest in patients’ daily lives. The development of the GPM-DAS was based on a synthesis of the three major conceptual models of GPM (*interpersonal hypersensitivity, intrapsychic coherence, and model of* overcontrol) and informed by our clinical experience with these frameworks. Examples were selected collaboratively with GPM experts, ensuring they reflected the core self-related and interpersonal features highlighted by these models. An initial pool of examples was generated, and the most representative examples were retained for each dilemma. We opted for a 10-point Likert scale, and the order of items on the scale was carefully considered. The thresholds for interpretation (0-2 (non-relevant), 3-5 (mildly relevant), 6-8 (moderately relevant), and 9-10 (strongly relevant)) were empirically and clinically determined to provide a nuanced understanding of the relevance of each dilemma. These thresholds will be further explored and validated within the context of this study, with future research aiming to refine their psychometric properties. These thresholds will also be used within the GPM-extended framework to help clinicians in prioritizing the most relevant dilemmas for each patient. It is important to note that the GPM-DAS was simultaneously developed in both French and English, ensuring broader applicability and facilitating its use across diverse clinical and research contexts. However, as this is a newly developed scale, it has not yet undergone formal validation, which will be addressed in future studies.

To assess personality functioning, we will use the LPFS-BF (34). The LPFS-BF has satisfactory internal consistency for total score (α = .69), and marginal to fair internal consistency for sub-scores (Self: α =.57; Interpersonal: α =.65). It is a 12-items scale used to assess personality functioning as described in the section III of the DSM-5 (21), with two sub-scales (self and interpersonal functioning), each item being rated on a 4-points Likert scale.

To assess personality traits, we will use the Personality Inventory for DSM-5—Brief Form (PID-5-BF) (45). The PID-5-BF is 25-items scale assessing the five personality traits of AMPD’s criterium B, (namely, negative affectivity, detachment, antagonism, disinhibition, and psychoticism), each item being ranked on a 4-points Likert scale, and has a satisfactory internal consistency (α ranging from .68 for antagonism to .81 for total score).

To assess self-reported borderline symptoms, we will use the Borderline Symptom List −23 items (BSL-23) (46). The BSL-23 assesses the global severity of BPD symptoms, each item being ranked on a 5-point Likert scale (from 0 (“never”) to 4 (“always”)) and has high internal consistency (α= 0.935-0.969).

To assess pathological narcissism, we will use the PNI-B (47,48). The PNI-B is a 28-items scale assessing the two main facets of pathological narcissism (grandiosity and vulnerability), each item being ranked on a 5-points Likert Scale, and has great internal consistency (for grandiosity subscale: α=.83-.86; for vulnerability: α= .93). To our knowledge, no thresholds are currently available to distinguish narcissistic patients from clinical controls using the PNI-B, but mean PNI-B grandiosity and vulnerability scores found in general population in the original validation study were between 2.7 and 2.9 for grandiosity, and between 1.9 and 2.5 for vulnerability (48).

Finally, to assess obsessive-compulsive symptoms, we will use the POPS (49,50). The POPS is a 49-items scale, each rated on a 6-point Likert scale, that assesses the overall severity and five facets of obsessive-compulsive personality (namely, difficulty with change, emotional over-control, rigidity, maladaptive perfectionism, and reluctance to delegate), with great internal consistency (for sub-scores: α=.89 to .93; for total score: α=.97) (49). To our knowledge, no thresholds are currently available to distinguish OCPD patients from clinical controls using the POPS, but the mean POPS total score found in general population in the original validation study was 163.13 (SD=36.60) (50).

##### Other self-report assessment

To assess emotion regulation difficulties, we will use the DERS, in its original 36-items version (51). The DERS assesses emotion regulation difficulties in 6 different domains, ranging from awareness to acceptance of emotions or to ability to access to effective emotion regulation strategies. Each item is rated a 5-point Likert scale. The original DERS has great internal consistency (for sub-scores: α = .80 to .89; for total score: α = .93).

To assess impulsivity, we will use the original UPPS scale (52,53). The UPPS is a 45-items scale assessing impulsivity on four dimensions (urgency, lack of premeditation and perseverance, and sensation seeking), each item being rated on 4-point Likert scale, and has good internal consistency (α from .77 to .83).

To assess entitlement, we will use the Psychological Entitlement Scale (PES) (54). The PES is a 9-items questionnaire assessing overall level of entitlement, each item being rated on a 7-points Likert scale, and has great internal consistency.

To assess alexithymia, we will use the 20-item version of the Toronto Alexithymia Scale (TAS-20, (55)). The TAS-20 assess alexithymia in three main dimensions (difficulty describing feelings, difficulty identifying feelings, externally oriented thinking), each item being rated on a 5-points Likert scale, and is a reliable and valid instrument with good psychometric properties (55)

To assess mentalizing abilities, we will use the Mentalization Scale (MentS) (56). The MentS scale is a 28-items scale assessing mentalizing abilities, with acceptable internal consistency in clinical samples (e.g., α = .75 for total score), and with three reliable subscales (self-mentalization, other-mentalization, and motivation to mentalize). Each item is rated on a 5-points Likert scale, and a total score can be calculated.

To assess non-suicidal self-injurious (NSSI) and suicidal behaviors, we will use the Suicidal Behaviors and Body Damage and Modifications Scale (SBBDM-S) (57), a 10-items self-report questionnaire recently developed (and currently not validated), that assesses the lifetime prevalence of suicidal behaviors, body modifications and NSSI.

To assess social functioning, we will use the Questionnaire de Fonctionnement Social (QFS) (58). The QFS is a 16-items questionnaire assessing the frequency and satisfaction of social behaviors, each item being rated on a 5-points Likert scale, and has good internal consistency (α = .65 - .83).

To assess disability and functional impairment, we will use the Sheehan Disability Scale (SDS) (59–61). In the SDS, the patient rates on a 10-point visual analog scale (graded by 5 levels described by words: not at all, slightly, moderately, very much, very much) the extent to which 3 domains of his/her life (work/school; social life and family life/family responsibilities) are impacted by his/her symptoms. The scores for the 3 domains can be summed to give an overall functional index ranging from 0 (no impairment) to 30 (severely impaired). The SDS has good psychometric properties, including internal consistency (e.g., for total score, α = .89).

To assess attachment styles, we will use the Experience in Close Relationship – Revised version (ECR-R) (62,63). The ECR-R is a 36-items questionnaire with good psychometric properties assessing attachment anxiety and avoidance, each item being rated on a 7-point Likert scale (62).

To assess defense mechanisms, we will use the Defense Mechanism Rating Scales Self-Report (DMRS-SR) (64). The DMRS-SR is a 30-items questionnaire assessing the whole hierarchy of defense mechanisms, each item being rated on a 5-point Likert scale, and has great internal consistency (α =.75 to .90).

Finally, to assess working alliance, we will use the Working Alliance Inventory – Short Form Revised (WAI-SR) (65). The WAI-SR is 12-items questionnaires investigating three domains of therapeutic alliance (agreement between patient and therapist on 1°) the goals of the treatment and 2°) about the tasks to achieve these goals, and 3°) the quality of the bond between the patient and therapist), each item being rated on a 5-points Likert scale. The WAI-SR has high internal consistency (e.g., for total score: α = .95).

#### Fidelity assessment

The fidelity of the individual GPM intervention will be assessed by two independent certified GPM supervisors that will assess 2 videos of interview from each therapist involved in the study, one recorded in the first months of therapy and one in the last months. The supervisors will rate the interview using a scale developed based on the General Psychiatric Management Adherence Scale guidelines (66).

### 2.7. Statistical analysis plan

All the analyses will be conducted using R and R studio (67). The study’s principal investigator (MB) is responsible for drafting and implementing the statistical analysis plan.

#### 2.7.1. Endpoints

The primary endpoint will be the difference in the overall intensity of BPD symptoms, as assessed by the ZAN-BPD, between the two groups after 1 year of treatment.

Secondary endpoints will be the difference in progression after 1 year of treatment on the various sub-scores of the ZAN-BPD (affective, cognitive, impulsive, interpersonal); the personality functioning, as measured by the LPFS-BF and its two sub-scores (self and interpersonal functioning); the intensity of the five personality traits of the alternative DSM-5 model, as measured by the PID-5-BF; the level of disability, as measured by the SDS and its three sub-scores (work/education, social life, family life); the level of social functioning, measured by the QFS and its two sub-scores (frequency and satisfaction); the number of suicide attempts and self-injurious behaviors, as measured by the SBBDMS; the self-reported intensity of borderline symptoms, as measured by the BSL-23; the intensity of narcissistic vulnerability and grandiosity, as measured by the PNI-B; the intensity of obsessive symptoms, as measured by the POPS and its five sub-scores (rigidity, excessive emotional control, inadequate perfectionism, reluctance to delegate, difficulty changing); the intensity of impulsivity, as measured by the UPPS and its four sub-scores (urgency, lack of perseverance, lack of premeditation, sensation seeking); the intensity emotional dysregulation, as measured by the DERS and its 6 sub-scores (lack of awareness of one’s emotions, lack of clarity about the nature of one’s emotions, lack of ability to engage in emotionally oriented activities); the intensity of alexithymia, as measured by the TAS-20 and its three sub-scores (description of feelings, identification of feelings, externally-oriented thoughts); the mentalizing abilities, as measured by the MentS and its three sub-scores (mentalizing self, mentalizing other, motivation to mentalize); the type of attachment styles, as measured by the ECR-R; the overall intensity of entitlement, as measured by the PES; the type of defense mechanisms, as measured by the DMRS-SR; the self-reported quality of therapeutic relationship, as measured by the WAI-SR and its sub-scores (goal, work, bond); the frequency of categorical personality disorder diagnoses, as measured by the SCID-II; and the frequency of current comorbidities, as measured by the MINI. In addition, differences in terms of partial response, complete response, and remission rates will be assessed, as will therapy discontinuation rates during the treatment year. To note, response and remission will be defined on the basis of the standards used by Ridolfi and colleagues (35) : a “partial response” is defined as a decrease of > 4 but < 8 points in the total score on the ZAN-BPD scale, a “response” is defined as a decrease of > 8 points, and a “remission” is defined as moving from one of the higher stages of intensity to “mild symptoms” (with 1 < score < 9 : mild symptoms; 10 < score < 18: moderate symptoms; 19 < score < +: severe symptoms).

#### 2.7.2. Headcount calculations and patients included in the analysis

On the basis of several recent randomized controlled trials using the ZAN-BPD score as the primary outcome (68–70) and using their expected clinically significant difference and standard deviation, we estimated that 64 patients per group should be included to have 80% power to detect a clinically significant difference of 4 points (SD=8) during the year of treatment, with a two-sided alpha risk of 5%. Assuming a lost to follow-up rate of around 20%, we estimated that a minimum of 77 patients should be included in each group. Finally, as the secondary analyses are exploratory, we have not planned any adjustment for multiple comparisons, and the statistical threshold retained is 0.05.

All patients having initiated the program and having initial data will be included in the analysis. If data are missing at 1 year, they will be imputed using multiple imputations. For this, we will use the R package “mice” (71). We also plan to perform sensitivity analyses, including an analysis of subjects with no missing data (“completers”).

#### 2.7.3. Analyses

For the primary endpoint, we will use a multiple linear regression model, with the mean ZAN-BPD score at 1 year as dependent variable, and with a binary variable (taking 0 for the “classic GPM” group, and 1 for the “GPM-extended” group) and the baseline ZAN-BPD scale score as independent variables. Indeed, we want to adjust our model on the baseline ZAN-BPD scale score given that the main prognostic factor for treatment response in BPD patients is baseline symptom intensity (72). To note, in sensitivity analysis, other adjustment variables will be added according to any inter-center differences detected on patient characteristics at baseline (prognostic factors).

We will reproduce the same approach for all the secondary endpoints, namely, a regression model, taking the 1-year score of the variable as the dependent variable, and a binary variable (taking 0 for the “GPM classic” group, and 1 for the “GPM-extended” group) and baseline score of the variable (when available) as independent variables. The model will be adapted according to the type of dependent variable: linear regression model for a quantitative judgment criterion, logistic model for a binary dependent variable, quasi-Poisson and negative binomial model for a count. Adjustment variables will be added if relevant for sensitivity analysis.

Finally, we do plan to carry out exploratory analyses (including mediation) using data from all patients included, regardless of their group. These analyses will not, however, include a study of the influence of the type of therapy on the patient’s evolution. Rather, the aim will be to assess the factors influencing patient improvement during a GPM-based treatment, independently of the adaptation. This will not be the subject of the present study but rather of other targeted studies.

## 3. Discussion

In this article, we described the protocol we elaborated to evaluate our new dimensional adaptation of GPM (GPM-extended). In this discussion, we want to underline the potential benefits of our research alongside its possible methodological flaws.

Indeed, the EPSYLIA project, if carried out as planned, will produce new data regarding the efficacy of GPM-based intervention to treat patients fulfilling BPD criteria. This is all the more relevant given the small amount of research conducted on GPM efficacy, especially on long-term treatment, and the potential of diffusion this model has on a wide scale (73). Indeed, to date, only 5 randomized controlled trials have been conducted, one comparing DBT and GPM after 1 year of treatment (30,31), two comparing a brief (10 sessions) individual version of GPM (still as a comparison treatment) to a personalized-treatment based on case formulation (74,75), one compared the same brief individual version to treatment as usual (76), and one comparing a 6-week GPM-based psychoeducation group to a waiting list (35). All these studies found interesting results on the actual efficacy of GPM to improve borderline symptoms, as well as self and interpersonal functioning. However, only McMain’s study assessed the efficacy of a long-term GPM treatment. Thus, despite being observational, the EPSYLIA study could add further evidence on the efficacy of a long-term GPM-based treatment to treat patients with BPD.

On the other hand, we believe that our adaptation and the EPSYLIA project could also be a first step in solving the current conflicts at play in the personality disorders field. Indeed, as described in the introduction, classification of personality disorders is undergoing a paradigm shift from traditional diagnostic categories to dimensional systems. If these dimensional models are thought to represent a more valid approach of personality pathology, there are concerns regarding the deletion of the BPD category. Indeed, a large amount of research has been conducted among people who meet categorical BPD criteria, and, without the diagnosis, some experts fear that patients would lose access to specialized treatments for this condition, notably given the absence of treatment developed based on dimensional models (77). By building an in-between model, incorporating both the positive aspects of BPD category (e.g., existence of evidence-based treatment programs) and of dimensional models (e.g., consideration of the primary importance of self- and interpersonal functioning in personality assessment beyond diagnostic categories, and of within-disorder heterogeneity and high rates of comorbidity), we believe that the EPSYLIA project could be one of the first adaptation to try to pragmatically solve this issue.

However, a significant number of potential methodological issues must be underlined and will be considered during the study. Despite limits inherent to the design of the study (e.g., observational, non-randomized study), one major risk of bias is that the primary investigator (MB) will have the triple role of therapist, trainer and supervisor in the “GPM-extended” group, in addition to double role of trainer and supervisor in the “classic GPM” group. The reasons for this multiplicity of roles are primarily organizational, as the primary investigator is a practicing psychiatrist in the structure where the “GPM-extended” group will be treated. We believe that there is an inherent risk of bias in being both therapist and trainer/supervisor, especially when considering that the primary investigator carries out this study aiming to show the superiority of GPM-extended over classic GPM. To mitigate this risk, we plan to have the “classic GPM” team complete the official online training of the Gunderson Personality Disorder Institute (78), an institute not directly involved in the study (even though Lois Choi-Kain act as a theoretical support in this study), alongside the training with the primary investigator. In addition, we plan to regularly involve independent GPM supervisors in the “classic GPM” team’s supervisions (approximately once every 1-2 months, depending on availability), to ensure that these supervisions are carried out properly. We believe that these two elements will reduce the risk of bias in the training and supervision of the “classic GPM” team. Another important limitation lies in the possibility that BPD criteria, while serving as a means to assess personality dysfunction (28), may also be inherently biased toward a specific subtype or dilemma characteristic of BPD. Consequently, the reliance on a BPD diagnosis made using the SCID as the primary inclusion criterion introduces a risk of selection bias, particularly with respect to the interpretation of future findings on the efficacy of GPM and/or GPM-extended in improving dimensional personality functioning. In other words, even if significant improvements in personality functioning are observed with GPM and/or GPM-extended, these results should be interpreted cautiously, notably because GPM is already established as an effective treatment for individuals. Finally, an additional source of bias may stem from the fact that participants in both study arms will be allowed to engage in other forms of general psychiatric care (e.g., trauma-focused or addiction-focused treatments), provided they do not target personality disorders. While this approach respects the ecological validity of real-world treatment settings, it also introduces the possibility that improvements in outcomes may partly reflect the influence of these ancillary interventions, rather than the study treatment alone. To mitigate this risk, we will try to limit and record such additional interventions as much as possible during the treatment year, notably to adjust for their potential influence, if needed. Despite these limitations, we believe that our study, if conducted rigorously, has the potential to contribute meaningfully to the existing literature on GPM. This is particularly notable given its long-term scope (1 year), its prospective controlled design, and its incorporation of a more dimensional approach to the assessment of personality disorders.

## Data Availability

No datasets were generated or analysed during the current study. All relevant data from this study will be made available upon study completion

## 4. Abbreviations

AMPD: Alternative Model of Personality Disorders
BPD: Borderline Personality Disorder
BSL-23: Borderline Symptom List – 23 Items
DBT: Dialectical Behavioral Therapy
DERS: Difficulties in Emotion Regulation Scale
DMRS-SR: Defense Mechanism Rating Scales Self-Report
ECR-R: Experience in Close Relationships – Revised
GPM-DAS: GPM Dilemma Assessment Scale
ICD-11: International Classification of Diseases, 11th Version
LPFS-BF: Level of Personality Functioning Scale - Brief Form
MBT: Mentalization-Based Therapy
MentS: Mentalization Scale
MINI: Mini-International Neuropsychiatric Interview 7th Version
NPD: Narcissistic Personality Disorder
OCPD: Obsessive-Compulsive Personality Disorder
PD: Personality Disorder
PES: Psychological Entitlement Scale
PID-5-BF: Personality Inventory for DSM-5—Brief Form
PNI-B: Pathological Narcissism Inventory - Brief Form
POPS: Pathological Obsessive-Compulsive Personality Scale
QFS: Questionnaire de Fonctionnement Social
SBBDM-S: Suicidal Behaviors and Body Damage and Modifications Scale
SCID-II: Structured Clinical Interview DSM–IV Axis II Personality Disorders
SDS: Sheehan Disability Scale
TAS-20: Toronto Alexithymia Scale – 20 Items
UPPS: UPPS-P Impulsive Behavior Scale
WAI-SR: Working Alliance Inventory – Short Form Revised
ZAN-BPD: Zanarini Rating Scale for Borderline Personality Disorder

## 5. Declarations

### Ethics approval and consent to participate

Not applicable.

### Consent for publication

Not applicable.

### Availability of data and materials

Not applicable.

### Authors’ contributions

MB was responsible for the construction of the study, wrote the first draft, and reviewed every version of the manuscript. IB, MZ, MA, AUM, FC, MLP, MB, PL, OL, and SL participated actively in the construction of the study and reviewed every version of the manuscript. LCK and MS supervised and participated actively in the construction of the study and reviewed every version of the manuscript. Every author approved the final version of the manuscript.

## Acknowledgements

None.

